# Genotype-phenotype discrepancy among family members carrying a novel glucokinase mutation: insights into the interplay of GCK-MODY and insulin resistance

**DOI:** 10.1101/2024.08.13.24311668

**Authors:** Shuhui Ji, Hua Shu, Hongqiang Zhao, Yuanyuan Ye, Xuan Liu, Shanshan Chen, Ying Yang, Wenli Feng, Jingting Qiao, Jinyang Zhen, Xiong Yang, Ziyue Zhang, Yu Fan, Yadi Huang, Qing He, Minxian Wang, Kunlin Wang, Ming Liu

## Abstract

**Aims/Hypothesis:** Heterozygous inactivating mutations in the glucokinase (GCK) gene are known to cause maturity-onset diabetes of the young (GCK-MODY). We identified a novel variant of uncertain significance (VUS) GCK mutation (c.77A>T, p.Q26L) in two family members presenting markedly different severities of diabetic phenotypes. This study aimed to elucidate the potential diabetogenic effect of GCK-Q26L and to explore the mono- and poly-genetic background attributing to different diabetes phenotypes.

**Methods:** Whole-exome sequencing (WES) and genetic analyses, including polygenic risk score (PRS) assessments, were performed in three members of a family with early-onset diabetes. To elucidate the impact of the GCK-Q26L mutation on glucose homeostasis, a global knock-in mouse model harboring this mutation in both heterozygous and homozygous states was generated. Insulin content and insulin secretion response to glucose and potassium were evaluated in isolated islets. Furthermore, the effects of dorzagliatin (a glucokinase activator, GKA) and liraglutide (a glucagon like peptide 1 receptor agonist, GLP-1RA) on glucose tolerance and insulin secretion were assessed in GCK-Q26L mutant mice.

**Results:** The proband, who inherited the GCK-Q26L mutation from her father (presenting with non-progressive, mildly elevated blood glucose), exhibited severe diabetic phenotypes including polydipsia, polyuria, polyphagia, weight loss, and ketosis, accompanied by significant dyslipidemia. Genetic analyses revealed that the proband’s severe phenotypes and metabolic profiles were associated with a high polygenic risk score (PRS) for insulin resistance that was inherited from her mother. Global heterozygous GCK-Q26L knock-in mice showed a mild increased fasting blood glucose, impaired glucose tolerance (IGT), and decreased serum insulin. Homozygous GCK-Q26L mice presented more severe phenotypes compared to their heterozygous counterparts, confirming the diabetogenic nature of the GCK-Q26L mutation. Further analyses indicated that GCK-Q26L did not affect insulin sensitivity and islet insulin content. However, GCK-Q26L blunted islet responsiveness to different glucose concentrations and markedly impaired glucose-stimulated insulin secretion (GSIS) without affecting potassium chloride-stimulated insulin secretion (KSIS) and glucose inhibitory effects on glucagon secretion. Both GKA and GLP-1RA enhanced insulin secretion and improved glucose tolerance in mutant mice.

**Conclusions/Interpretation:** This study demonstrates that GCK-Q26L is a GCK-MODY causing mutation. The interplay of GCK-Q26L with a high PRS for insulin resistance contributes to severe diabetic phenotypes. The findings not only expend the list of GCK-MODY causing mutations originally classified as VUS mutations, but also provides insights into interactions of GCK-MODY with polygenic risks of type 2 diabetes, highlighting the importance of considering polygenic backgrounds in the assessment and management of monogenic diabetes.

**Research in Context:** What is already known about this subject?

Heterozygous inactivating mutations in the GCK gene cause GCK-MODY, an autosomal dominant disorder characterized by mild hyperglycemia present from birth.

Insulin resistance can be influenced by multiple genetic polymorphisms, contributing to varying diabetes phenotypes.

What is the key question?

Is the newly discovered GCK mutation pathogenic?

Do the interactions between the GCK mutation and PRS for insulin resistance influence the phenotypic variability in patients carrying GCK-MODY?

What are the new findings?

The study demonstrates GCK-Q26L impairs GSIS and causes diabetes, establishing it as a novel GCK-MODY causing mutation originally classified as a VUS mutation.

The GCK-Q26L knock**-**in mouse line replicates phenotypes of GCK-MODY in humans, establishing it as an excellent model for GCK-MODY.

The phenotypic variability in patients with GCK-MODY can be significantly influenced by high-risk genetic predisposition of type 2 diabetes.

Both GKA and GLP-1RA enhance insulin secretion and improve glucose tolerance in GCK-Q26L mutant mice, suggesting that they are favorite options for treatment of patients with GCK-MODY and insulin resistance.

How might this impact clinical practice in the foreseeable future?

Recognizing atypical presentations of monogenic diabetes influenced by polygenic factors can enhance diagnostic accuracy and personalized management.

Genetic testing and polygenic risk score assessments can help identify patients at higher risk of severe phenotypes, allowing for earlier and more targeted interventions.

## Background

Glucokinase (GCK), a key enzyme in the glycolytic pathway, plays a critical role in glucose metabolism. It acts as a glucose sensor in pancreatic β cells and regulates insulin secretion in response to blood glucose levels [1, 2]. The unique kinetic properties of GCK make it ideal for this regulatory role, as it responds to even small changes in glucose concentration within the physiological range [2, 3]. Heterozygous inactivation mutations of GCK are known to cause maturity-onset diabetes of the young type 2 (MODY 2, also known as GCK-MODY), an autosomal dominant form of diabetes. These GCK inactivation mutations impair enzymatic activity, affecting glucose sensing of pancreatic β cells, resulting in a higher set point for insulin secretion, and therefore leading to mild and non-progressive elevated fasting blood glucose (FBG) from an early age [4, 5]. Notably, unlike other types of diabetes, GCK-MODY does not typically increase the risks of chronic micro- and macrovascular complications of diabetes [6]. Therefore, the current recommendations for the management of GCK-MODY emphasize lifestyle management and regular blood glucose monitoring rather than active treatment with hypoglycemic agents [6, 7]. However, the clinical presentation of GCK-MODY can vary significantly among individuals, even within the same family. The underlying mechanisms for this variability remain incompletely understood.

Insulin resistance associated with multiple genetic and environmental factors is a key feature of type 2 diabetes (T2D) [8]. Recent studies have identified various genetic polymorphisms contributing to insulin resistance, each adding a small risk but cumulatively having a significant impact [9]. A high polygenic risk score (PRS) for insulin resistance aggregates the effect of numerous risk alleles, increasing the likelihood of T2D development [10, 11]. Additionally, a high PRS for T2D may also interact with monogenic diabetes causing mutations, leading to atypical and severe diabetic phenotypes that challenge the traditional approaches for classification and management of different types of diabetes [8, 12–14]. Understanding the interaction between these genetic factors is crucial for developing personalized treatment strategies [15].

In this study, we identified a novel variant of uncertain significance (VUS) GCK mutation (c.77A>T, p.Q26L) in two members of a family exhibiting markedly different severities of diabetic phenotypes. A global knock-in mouse model established that GCK-Q26L impaired insulin secretion of β cells response to glucose and caused an increase of FBG and an impairment of glucose tolerance, thereby confirming this mutation is a GCK-MODY causing mutation. Genetic analyses revealed the interplay between the GCK-MODY mutation and a high PRS for insulin resistance contributed to the severe diabetic phenotypes in the proband carrying GCK-Q26L. The study not only expends the list of GCK-MODY causing mutations originally classified as VUS mutations, but also highlights genetic bases underlying phenotype heterogeneities of patients with monogenic diabetes.

## Materials and Methods

### Patients

The clinical information of the proband and her family members was detailed in Table 1, collected and documented in Tianjin Medical University General Hospital. Informed consent was obtained from all family members. The study was approved by the Tianjin Medical University General Hospital Ethics Committee (IRB2017-047-01).

**Table 1.**
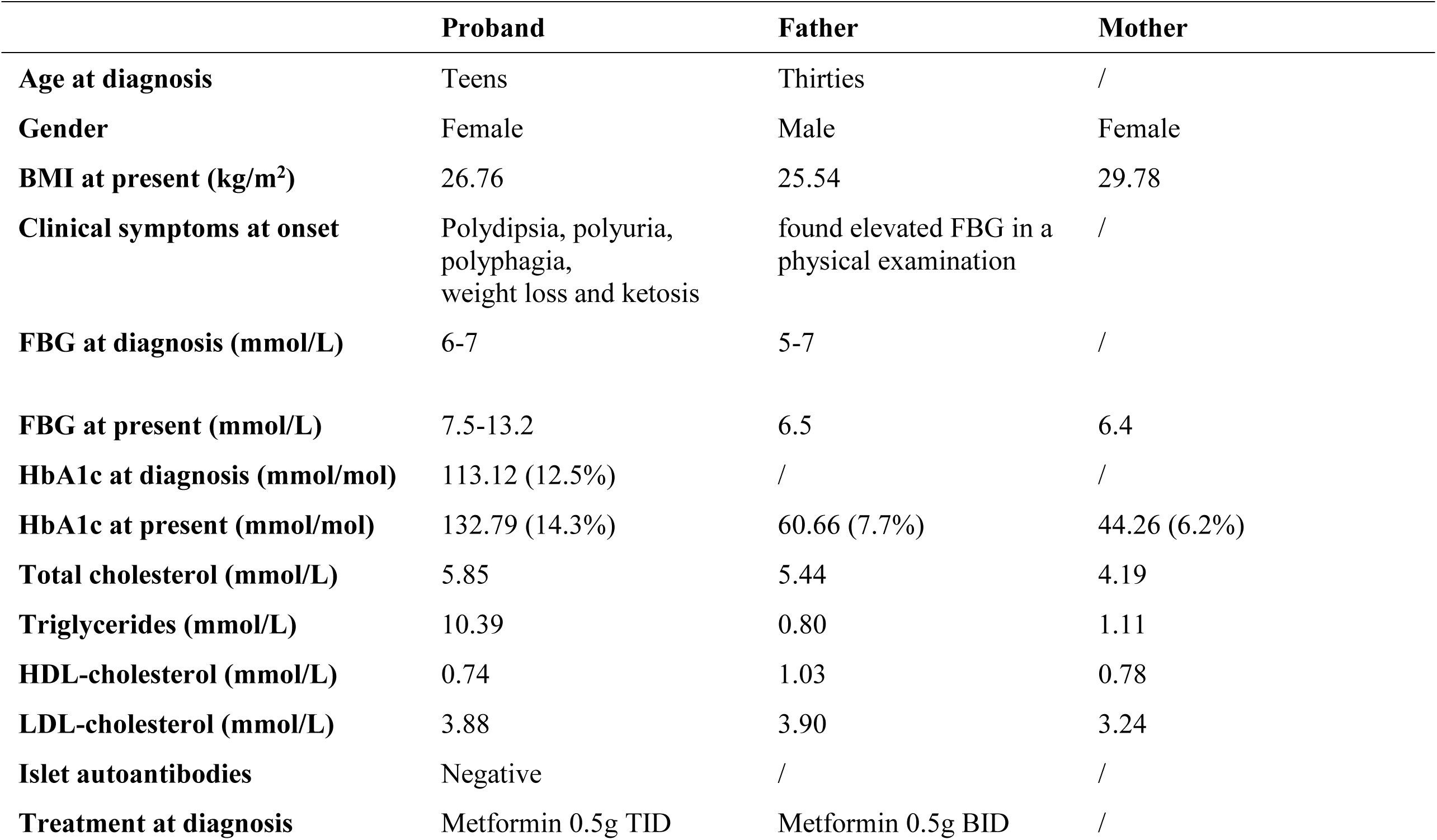
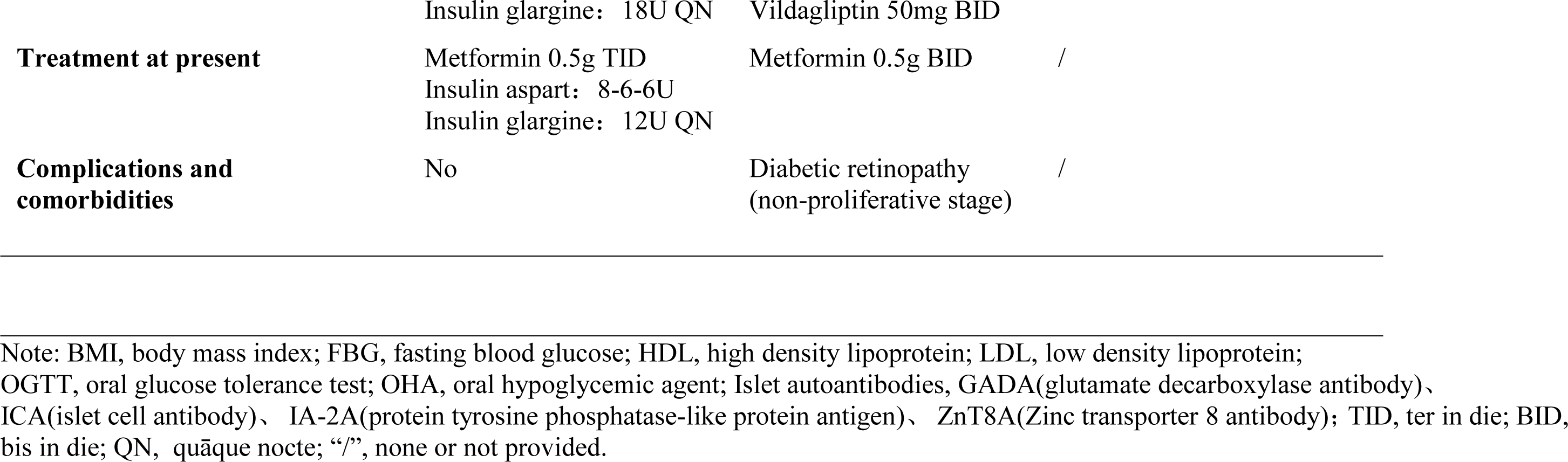
Clinical features of the family of the proband carrying GCK(Q26L) mutation.

### Genetic Testing and Analyses

DNA was extracted from proband’s white blood cells for targeted gene capture, followed by whole-exome sequencing (WES) utilizing massive parallel next-generation sequencing (NGS). The identified GCK gene mutation was further confirmed by Sanger sequencing, the primers for the forward- and reverse-strand were 5’-TGTGGGGGAGATGCCCCG-3’ and 5’-ACCAGAGGAGCCAAGGGTGAG-3’, respectively. Members of the proband’s family (father, mother and grandfather) were recruited and checked for the mutation using Sanger sequencing. Furthermore, the family members (father and mother) also underwent WES sequencing.

### Definitions for Insulin Resistance

Insulin resistance (IR) was measured as the ratio of triglycerides (TG) to high-density lipoprotein cholesterol (HDL-C) as described elsewhere [9, 16–19] (ESM Fig. 4). To correct the potential effect of cholesterol lowering medications, we adjusted the levels of TG and HDL-C before calculating IR with detailed adjustment in ESM Table 4.

### Genotype quality control and imputation

The proband trios were genotyped by Affymetrix Axiom whole-genome Asian Screening Array (ASA Array) with calling rate > 99%. Kingship (ESM Table1) and sex (ESM Table2) were further validated with genetically inferred ones. The data was merged with non-close related samples from the 1000 Genomes Project with ambiguous variants (A/T or G/C alleles) removed. This was followed by a series of quality controls in samples with a missingness rate > 5%, or heterozygosity out of 5 standard deviations were excluded, and variants with a missing rate > 1% or Hardy-Weinberg equilibrium with a P-value < 1 × 10^−6 were excluded. Then, principal component analysis (PCA) was conducted, and ancestry was inferred by the PCA clusters. Finally, the genotype was further imputed on the TOPMed imputation server [20] (ESM Fig. 3), and variants with imputation quality score (INFO) < 0.8 were removed in subsequent analysis.

### PRS assessment

We directly utilized the polygenic scores model for insulin resistance developed by Lotta et al.[8] from the PGS Catalog and calculated the PRS for our datasets together with 1000 Genomes Project East Asian samples by PLINK 2.0 [21]. To confirm the association between PRS and IR, we also calculated the PRS for the samples (424,068 passed quality control) from the UK Biobank (ESM Table 3) under application number 89885, following the same procedures. The PRS was further corrected by the value of principal components (PCs) for potential population structure and batch effects and standardized to normal distribution for further statistical analysis.

### Mice

We commissioned GemPharmatech Co., Ltd. to create a global knock-in GCK-Q26L mouse model. The Q26L site-directed mutagenesis was introduced in exon 2 of mouse GCK-201, changing the codon for the 26th amino acid from CAG to CTG and inserting MYC before the stop codon (ESM. Fig.1a). All mice were bred on a C57BL/6J background and housed in temperature (22–25 °C) and humidity (55 ± 5%) controlled rooms with a 12-hour light/dark cycle. All animal experimental protocols were approved by the Internal Animal Welfare Committee at Chu Hsien-I Memorial Hospital (Metabolic Diseases Hospital) of Tianjin Medical University.

### Glucose and insulin tolerance

Mice were fasted for 6 hours before intraperitoneal glucose tolerance tests (IPGTT). Glucose was administered via intraperitoneal injection at 2 g/kg body weight. Blood glucose was measured from the tail vein at 0, 15, 30, 60, and 120 minutes using an automated glucose meter (Roche, ACCU-CHEK, Germany). ELISA kits were used to measure plasma insulin (EZassay, China), and glucagon (Ruixin, China). For the intraperitoneal insulin tolerance test (IPITT), after a 4-hour fast, mice received an intraperitoneal injection of 0.7 U/kg body weight of insulin (Aspart, Novo Nordisk A/S, Denmark). Dorzagliatin (Desano, China) and Liraglutide (Solarbio, China) were administered to mice by gavage or intraperitoneal injection, respectively, 90 minutes before the start of the IPGTT.

### Measurement of insulin and glucagon secretion

Islet isolation, glucose-stimulated insulin secretion (GSIS), and potassium-stimulated insulin secretion (KSIS) were performed as previously described [22], glucose-inhibited glucagon secretion was performed simultaneously with GSIS. For the dose-dependent effects of glucose on insulin secretion, isolated pancreatic islets were sequentially cultured in Krebs-Ringer bicarbonate HEPES (KRBH) buffer with glucose concentrations of 2.8 mM, 3.3 mM, 4.5 mM, 5.6 mM, and 7 mM for 2 hours each. After incubation, supernatants were collected, and the islets were homogenized. Secreted insulin and islet insulin content were measured using ELISA kits (EZassay, China). The secretion efficiency was calculated with secreted insulin in the media normalized to islet insulin content.

### Islet perfusion assays

Islet perfusion assays were performed as previously described [22]. The liquid input flow rate was 150 µl/min, with samples collected every minute. Insulin secretion at each time point was quantified using the Wide Range Insulin Assay Kit (EZassay, China) according to the manufacturer’s instructions.

### Western blot analysis

Equal amounts of islet proteins were loaded onto 4-12% NuPage gradient gels (Thermo, USA), while other tissues were loaded onto 12.5% gels (US EVERBRIGHT, China). The antibodies used in this study were as follows: anti-GCK (Cat: A00884-1, BOSTER, China), anti-c-MYC (Cat: RMYC-45A-2, ICL, USA), anti-GAPDH (Cat: AC033, Abclonal, China), anti-insulin (homemade).

### Immunofluorescence

Mouse pancreas was fixed, sectioned, and stained with different primary antibodies followed by Alexa-Fluor-conjugated secondary antibodies. The antibodies used in this study were as follows: anti-insulin (homemade), anti-glucagon (Cat: G2654, Sigma, USA), anti-somatostatin (Cat: ab30788, abcam, USA). Fluorescent images were visualized using an Axio Imager M2 microscope (Carl Zeiss, Germany).

### Statistics

All experiments were performed independently at least three times. Data are presented as means ± SEM. Statistical analyses were conducted using GraphPad Prism 8 software. An unpaired Student’s t-test was used for data analysis, with a threshold of P < 0.05 to declare statistical significance.

## Results

### Identification of a novel variant of uncertain significance (VUS) GCK mutation in a family with early onset diabetes

In the clinic, we observed a special case of diabetes. The female proband was diagnosed with diabetes at age teens, presenting with polydipsia, polyuria, polyphagia, weight loss, and ketosis. Her body mass index (BMI) was 22.66 kg/m², and islet autoantibodies were all negative. Initial FBG was 6–7 mmol/L, 2hBG was 15–18 mmol/L, and HbA1c was 113.12 mmol/mol (12.5%). The result of the 75g oral glucose tolerance test (OGTT) confirmed the diagnosis of diabetes and indicated the presence of insulin resistance and inadequate insulin secretion in response to glucose load. (Fig. 1a). She was treated with Metformin (0.5 g TID) and Insulin glargine (18 U QN), with occasional monitoring showing FBG of 5– 7 mmol/L and 2hBG of 13–14 mmol/L. She continued this treatment inconsistently, without dietary control, regular exercise, or routine SMBG. After 2.5 years, she discontinued insulin and continued Metformin (0.5 g TID), with occasional FBG readings of 6–7 mmol/L. Five years post-diagnosis, her FBG increased to 7.5–13.2 mmol/L, HbA1c to 132.79 mmol/mol (14.3%), and BMI to 26.76 kg/m². Her regimen was changed to Metformin (0.5 g TID), Insulin aspart (8-6-6 U before meals), and Insulin glargine (12–16 U QN), but glycemic control remained poor, with FBG of 7–10 mmol/L. Her father was diagnosed with diabetes at thirties, treated with Metformin 0.5 g BID and Vildagliptin 50 mg BID, with FBG of 5–7 mmol/L. Seven years later, his regimen was changed to Metformin 0.5 g TID and occasional Dapagliflozin, maintaining FBG at 6.5 mmol/L and HbA1c at 60.66 mmol/mol (7.7%). Her grandfather was diagnosed with diabetes at fifties, managed with oral hypoglycemic agents. Her mother was found to have pre-diabetes at forties based on blood glucose and HbA1c levels. Clinical details of the proband and her family were summarized in Table 1.

**Figure 1.**
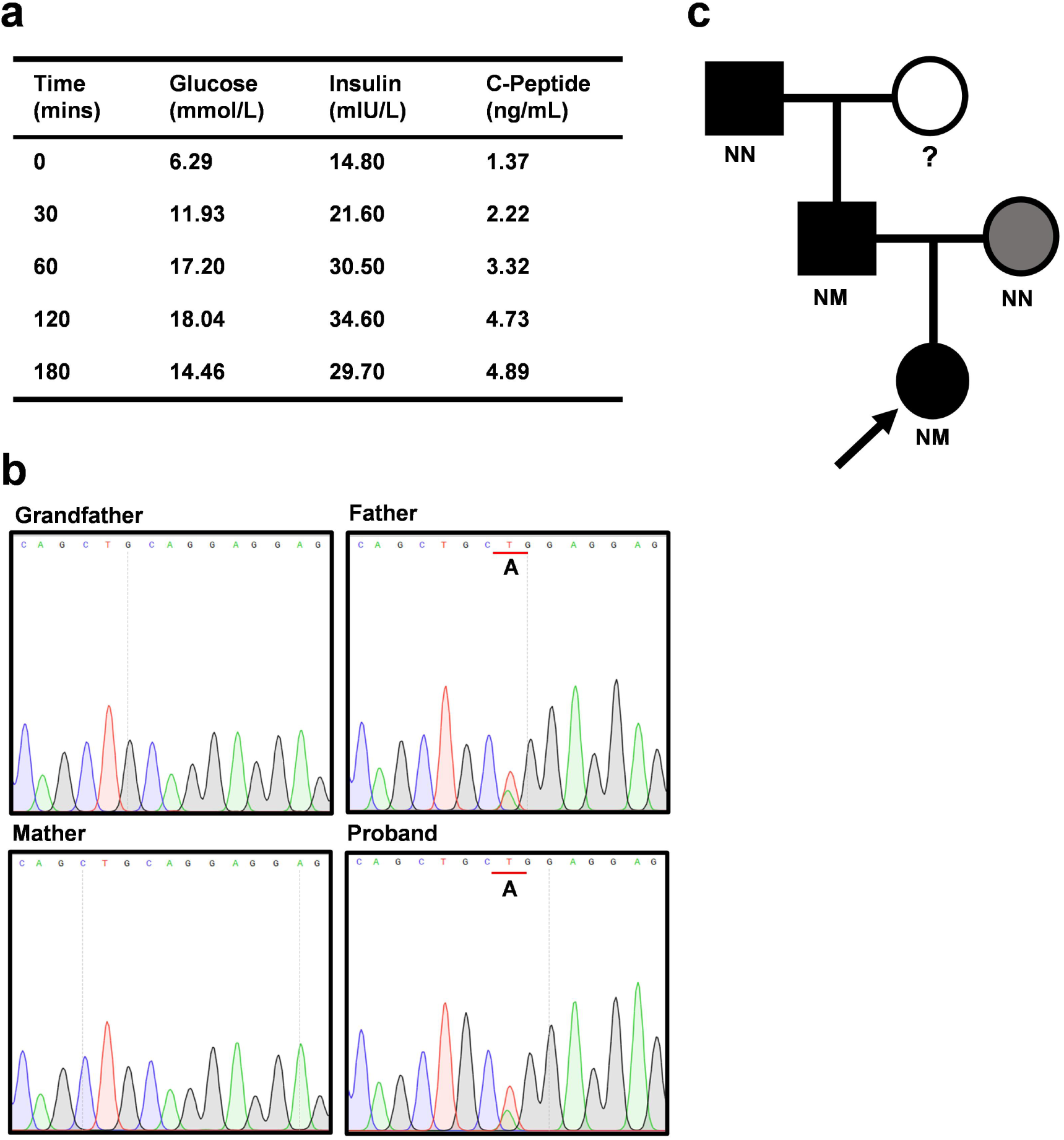
Proband with impaired insulin secretion and diabetes carries a GCK missense mutation (c.77A>T, p.Q26L) that was inherited from her father. (**a**) The results of blood glucose, insulin, and C-Peptide of the proband during OGTT performed at the onset of diabetes. (**b**) A missense mutation in GCK gene (c.77A>T, p.Q26L) was identified in the proband and her father by WES confirmed by Sanger sequencing. (**c**) Pedigree analyses showed that GCK-Q26L was inherited from proband’s father. Squares represent males and circles represent females. Filled black symbols indicated members affected with diabetes, gray symbol represented the member with the pre-diabetes. The proband was indicated with a black arrow. Where tested, the genotypes of family members were given (N, normal allele; M, mutant allele; NN, normal genotype; NM, heterozygous genotype).

Given the proband’s negative islet autoantibodies and family history, we suspected monogenic diabetes and performed genetic screening. DNA sequencing revealed a novel heterozygous missense mutation, c.77A>T, in exon 2 of the GCK gene, resulting in a glutamine-to-leucine substitution at position 26 (p.Q26L), which had not been reported yet. According to American College of Medical Genetics and Genomics (ACMG) and the Association for Molecular Pathology (AMP) in 2015 guidelines, the mutation was classified as a variant of uncertain significance (VUS) with evidence PM1+PM2+PP2. The biological information analysis software SIFT (http://provean.jcvi.org/index.php) predicts that the heterozygous variation c.77A>T (p.Q26L) in the GCK gene is deleterious. PolyPhen2 (http://genetics.bwh.harvard.edu) predicts a score of 0.026, indicating that the GCK gene mutation c.77A>T (p.Q26L) is benign. Additionally, the frequency of this mutation in the general (normoglycemic) population according to gnomAD (v4.1.0) is 0. Pedigree analysis showed that the proband’s father had the same heterozygous mutation, while the grandfather and mother did not (Fig. 1b, c).

Based on these findings, the proband’s diabetes type and the role of the GCK-Q26L mutation in her complex diabetic phenotype remain unclear. However, the father’s glycemic profile aligned with typical GCK-MODY characteristics. Therefore, further experiments were conducted to assess the impact of the GCK mutation on protein function.

### GCK-Q26L is a GCK-MODY causing mutation

Since GCK-Q26L was classified as a VUS and the proband presented atypical clinical features of GCK-MODY, it remained to be determined whether GCK-Q26L is a diabetes causing mutation. We therefore established a global knock-in mouse model expressing MYC-tagged GCK-Q26L (ESM Fig. 1a). Western blot confirmed the specific expression of MYC-tagged mutant GCK in the liver and islets in a dose-dependent manner in GCK-Q26L heterozygous (Het) and homozygous (Hom) mice (ESM Fig. 1b). We had monitored body weight (BW) and FBG of mutant and wild-type (WT) male mice for 16 weeks. Although the GCK-Q26L mutation did not affect BW, it caused mild, non-progressive elevation of FBG in Het mice, with further elevation observed in Hom mice (Fig. 2a, b). To investigate whether abnormal circulating levels of insulin or glucagon were associated with elevated FBG, we examined fasting serum insulin and glucagon levels. Our findings showed that although glucagon levels were unaffected, serum insulin levels were decreased in 16-week-old mutant mice (Fig. 2c, d). We therefore performed intraperitoneal glucose tolerance test (IPGTT) and found that Het and Hom mice exhibited significantly impaired glucose tolerance and lower serum insulin levels compared to that of WT mice at 3 weeks of age (Fig. 2e, f). The severities of decreased serum insulin and impaired glucose tolerance did not further worsen in 16-week-old mutant mice (Fig. 2g, h). The phenotype of female mice was similar to that of male mice but less severe (ESM Fig. 2). These data established that GCK-Q26L caused non-progressive mild hyperglycemia representing one of classic clinical features of GCK-MODY, confirming that GCK-Q26L is a GCK-MODY mutation.

**Figure 2.**
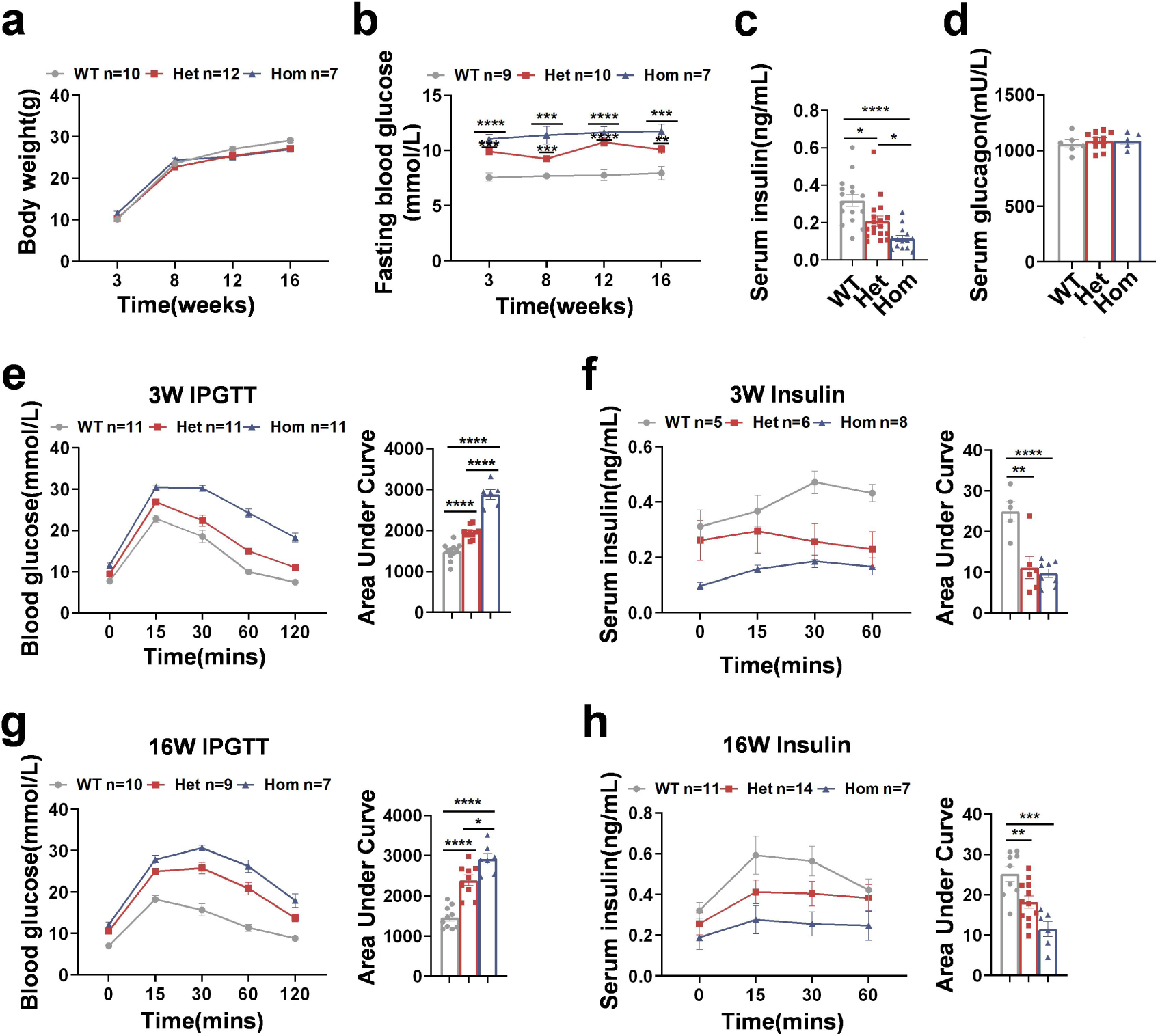
GCK-Q26L causes glucose intolerance in a dose-dependent manner. (**a**, **b**) Body weight (**a**) and fasting blood glucose (**b**) of male mice were measured at 3, 8, 12, 16 weeks of age. WT *vs* Het, WT *vs* Hom, ** P < 0.01, *** P < 0.001, **** P < 0.0001. (**c**) Fasting serum insulin in three-group 16-week-old male mice were measured by ELISA. (**d**) Fasting serum glucagon in three-group 16-week-old male mice were measured by ELISA. (**e**) IPGTT in 3-week-old male mice (2 g/kg i.p.) and the area under curve (AUC) of blood glucose during IPGTT. (**f**) Serum insulin levels in 3-week-old male mice during IPGTT were measured by ELISA, with calculated AUCs. (**g**) IPGTT in 16-week-old male mice (2 g/kg i.p) and the AUC of blood glucose. (**h**) Serum insulin levels in 16-week-old male mice during IPGTT were measured by ELISA, with calculated AUCs. Each data point represents an individual mouse. Values are shown as mean ± SEM. WT *vs* Het, WT *vs* Hom, Het *vs* Hom, *P < 0.05, **** P < 0.0001.

### GCK-Q26L does not affect insulin sensitivity and islet insulin content

Since GCK is also expressed in hepatocytes, we asked whether GCK-Q26L affected insulin sensitivity. We performed intraperitoneal insulin tolerance test (IPITT) and found that GCK mutant did not affect insulin sensitivity (Fig. 3a, b), suggesting that hyperglycemia was mainly due to decreased circulating insulin. To investigate whether reduced serum insulin in Het and Hom male mice is caused by decreased to islet insulin content or insulin secretion, we measured insulin content by two independent approaches (western blots and insulin enzyme-linked immunosorbent assays, ELISA). Both methods consistently showed no substantial differences in insulin content in islets of Het and Hom mice compared to WT mice (Fig. 3c-f). The ratio of proinsulin-to-insulin was also not affected (Fig. 3f). Immunofluorescence analysis revealed no impact of the mutation on the abundance and intraislet composition of insulin-positive, glucagon-positive, and somatostatin-positive cells among three groups (Fig. 3h). Furthermore, western blot results showed no significant differences in endogenous GCK protein expression in the islets among the three groups, indicating that the GCK-Q26L did not affect protein levels of endogenous GCK (Fig. 3d, g).

**Figure 3.**
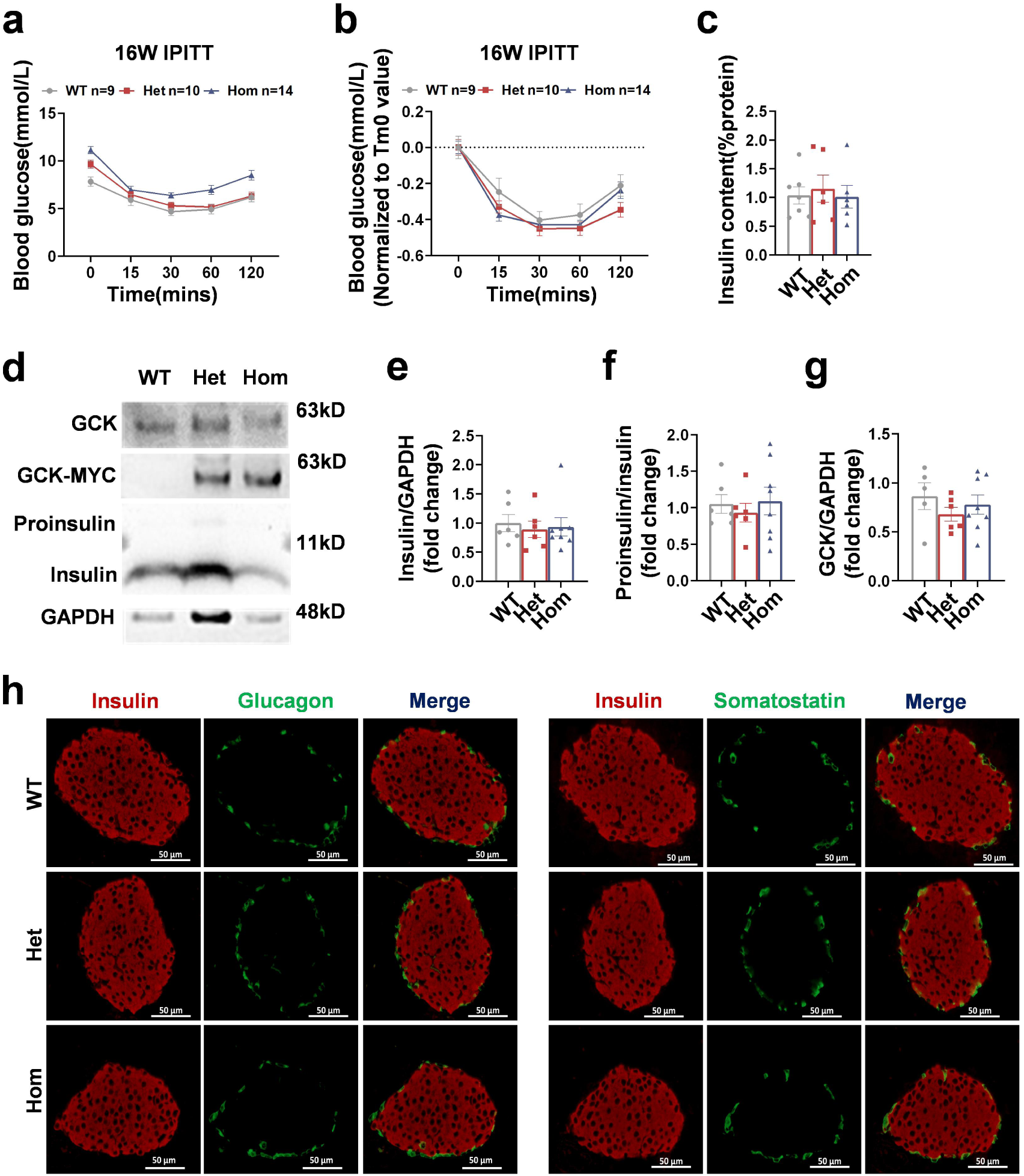
GCK-Q26L did not affect insulin sensitivity, insulin content, and islet cell composition. (**a, b**) IPITT were performed in 16-week-old male mice (0.7 U/kg i.p), (**b**) was (**a**) normalized to the baseline of blood glucose. (**c**) Islet insulin content of 16-week-old male mice was measured by ELISA. (**d**) Islets isolated from 10-week-old male mice were directly lysed. Western blots were performed to detect endogenous GCK, MYC-tagged GCK (GCK-MYC), proinsulin and insulin as indicated. GAPDH was used as a loading control. Quantification of insulin (**e**), the ratio of proinsulin to insulin (**f**), and GCK (**g**). (**h**) Representative immunofluorescence images of anti-insulin (red), anti-glucagon (green) and anti-somatostatin(green) antibodies in pancreatic sections from 12-week-old male mice. Scale bar = 50 μm (n = 3 mice/group). Each data point represents an individual mouse. Values were shown as mean ± SEM.

### GCK-Q26L impairs glucose-stimulated insulin secretion without affecting potassium-stimulated insulin secretion and glucose-inhibited glucagon secretion

Since the GCK-Q26L caused a decrease of circulating insulin without affecting islet insulin content (Fig. 2-3), we next asked whether it impair insulin secretion. Insulin secretion was monitored in isolated islets exposed to different glucose concentrations (2.8 mM, 3.3 mM, 4.5 mM, 5.6 mM, 7 mM). We found that the curve of glucose concentration-dependent insulin secretion was shifted to right in Het mice (Fig. 4a), suggesting that the glucose threshold for insulin secretion was increased in GCK-Q26L mice. In addition, insulin secretion response to even higher glucose (16.7 mM) was also impaired in Het islets and the response was further diminished in Hom islets (Fig. 4b). Islet perfusion tests with different glucose concentrations further supported these observations (Fig. 4c). However, despite defects in response to glucose, GCK-Q26L did not affect insulin secretion response to potassium in both tests of potassium chloride-stimulated insulin secretion (KSIS) and islet perfusion (Fig. 4d, e). Furthermore, no differences were observed in glucose-inhibited glucagon secretion (GIGS) in Het and Hom mutant mice (Fig. 4f). These data demonstrated that GCK-Q26L impaired the function of GCK as a glucose sensor in β cells, impeding GSIS without affecting KSIS and GIGS.

**Figure 4.**
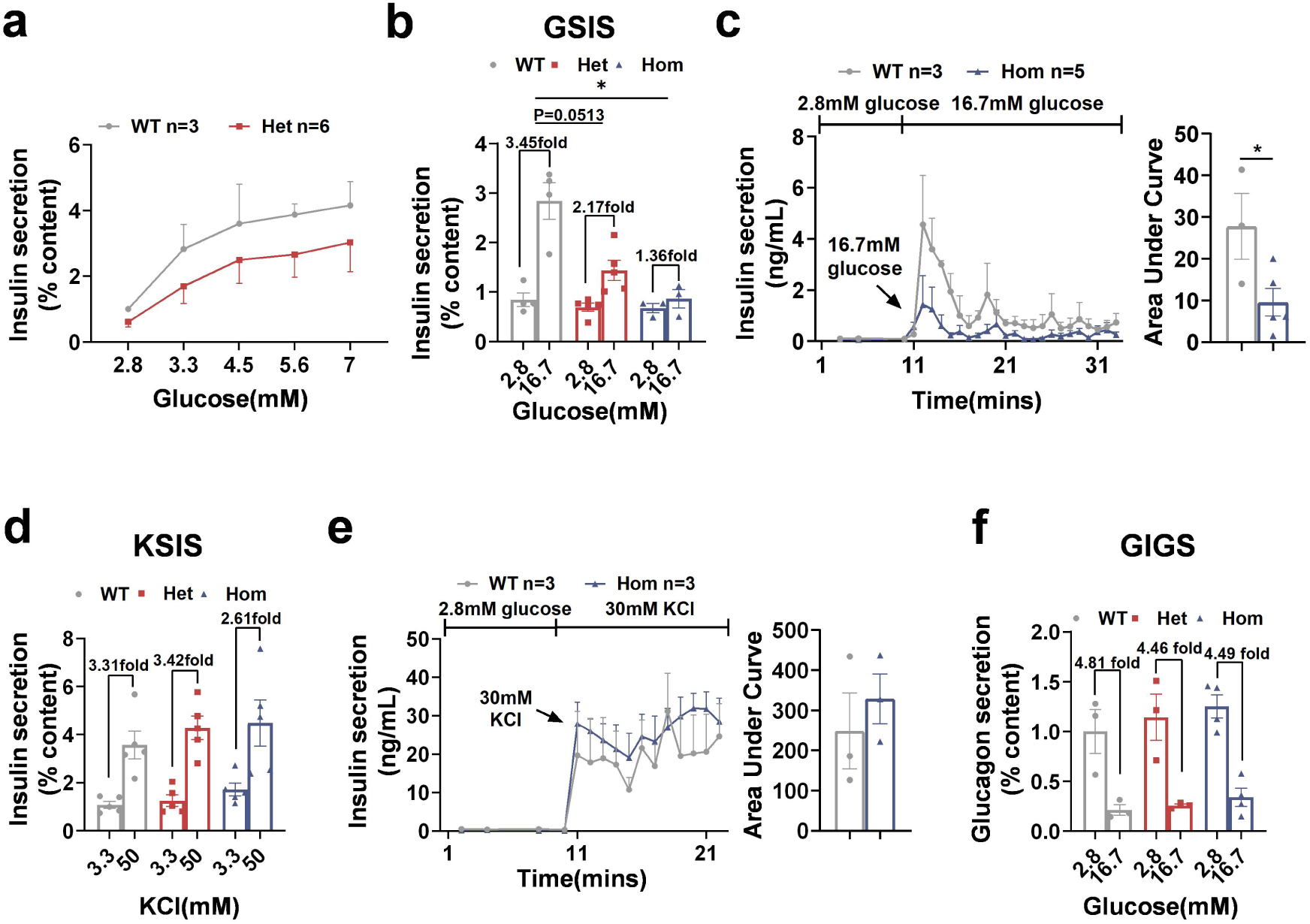
GCK-Q26L impaired glucose-stimulated insulin secretion without affecting potassium-stimulated insulin secretion and glucose-inhibited glucagon secretion. (**a**) Dose-dependent effects of glucose on insulin secretion of 16-week-old male WT and Het mice. Isolated islets successively incubated in 2.8, 3.3, 4.5, 5.6 and 7 mmol/L glucose. (**b**) Islets isolated from 10-week-old male WT, Het and Hom mice were incubated with 2.8 mmol/L for 1 hour followed by stimulated with either low (2.8 mmol/L) or high (16.7 mmol/L) glucose for 2 hours. Insulin secretion response to low and high glucose were measured. (**c, e**) Dynamic insulin secretion of 10-week-old male WT and Hom mice islets cultured in the perfusion chamber with continues buffer renewal (2.8 mmol/L glucose, 16.7 mmol/L glucose, 2.8 mmol/L glucose, 30 mmol/L potassium), with calculated AUCs. (**d**) Islets isolated from 10-week-old male WT, Het and Hom mice were incubated with 2.8 mmol/L glucose for 1 hour followed by stimulated with either low (3.3 mmol/L) or high (50 mmol/L) potassium for 2 hours. Insulin secretion response to low and high potassium were measured. (**f**) Glucagon secretion measured in isolated islets from 10-week-old male WT, Het and Hom mice when incubated in 2.8 and 16.7 mmol/L glucose. Each data point represents an individual mouse. Values are shown as mean ± SEM. * P < 0.05.

### Dorzagliatin restores the glucose threshold for insulin secretion and ameliorates glucose tolerance in GCK-Q26L mice

Dorzagliatin, a novel allosteric glucokinase activator (GKA), improved glycaemic control and was approved in China for the treatment of adult patients with type 2 diabetes [23, 24]. To evaluate the effects of dorzagliatin on insulin secretion and glucose tolerance in GCK-Q26L mice, we firstly treated mice with oral administrations of dorzagliatin, measured fasting blood glucose and insulin levels 90 minutes after treatment, and then performed IPGTT. We found that dorzagliatin significantly decreased FBG and increased fasting insulin levels in both WT, Het, and Hom mutant mice (Fig. 5a, b). IPGTT results further confirmed improvement of glucose tolerance in mutant mice treated with dorzagliatin (Fig. 5c). We further analyzed insulin secretion response glucose and found that dorzagliatin restored the glucose threshold for insulin secretion (Fig. 5d) and improved GSIS (Fig. 5e) in GCK-Q26L mutant islets.

**Figure 5.**
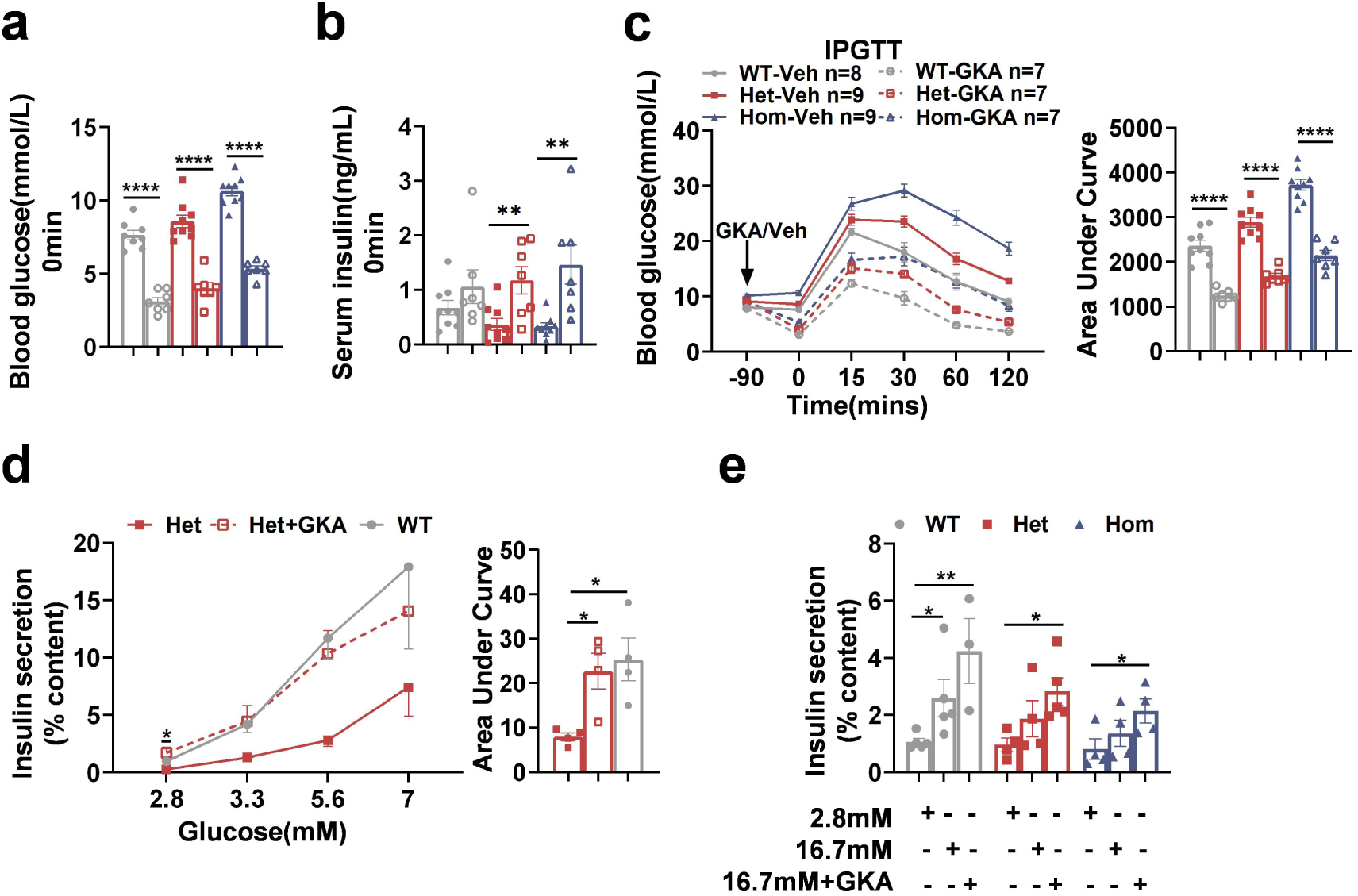
Dorzagliatin (GKA) restores the glucose threshold for insulin secretion and ameliorates glucose tolerance in GCK-Q26L mice. (**a-c**) We performed IPGTT on 10-week-old male mice (2 g/kg i.p), and vehicle (0.9% Nacl) or GKA (30 mg/kg) was administered orally to mice 90 minutes before glucose challenge. Blood glucose (**a**) and serum insulin level (**b**) before glucose challenge. (**c**) Blood glucose and AUC of IPGTT. (**d**) Dose-dependent effects of glucose on insulin secretion of 10-week-old male WT and Het mice with or without 30 uM GKA, and the AUC of Insulin secretion. Het *vs* Het-GKA, *P < 0.05. (**e**) Insulin secretion measured in isolated islets from 10-week-old male WT, Het and Hom mice when incubated in 2.8, 16.7 mmol/L glucose and 16.7 mmol/L glucose containing 30 uM GKA. Each data point represents an individual mouse. Values are shown as mean ± SEM. * P < 0.05, ** P < 0.01, **** P < 0.0001.

### Liraglutide amplifies insulin secretion response to glucose and ameliorates glucose tolerance in GCK-Q26L mice

Since liraglutide, a glucagon-like peptide-1 receptor agonist (GLP-1RA), has an effect of glucose-dependent stimulated insulin secretion, we next asked whether liraglutide could show beneficial effects on insulin secretion and glucose tolerance in GCK-Q26L mice. We conducted similar experiments as dorzagliatin treatment shown in Fig. 5. We found that unlike dorzagliatin, liraglutide did not significantly stimulate insulin secretion in the fasting state both in WT and mutant mice, and only moderately decreased FBG in GCK mutant mice compared with dorazgliatin (Fig. 6a, b). However, liraglutide indeed markedly stimulated insulin secretion and decreased blood glucose 15 minutes after glucose load (Fig. 6c, d), ameliorating overall glucose tolerance (Fig. 6e). In vitro experiments using isolated islets further confirmed that liraglutide improved GSIS (Fig. 6f) and amplified insulin secreation in 5.6 mmol/L glucose (Fig. 6g) in islets from GCK-Q26L mice. Altogether, these results suggest that while both dorzagliatin and liraglutide improve glucose homeostasis, they do so through distinct mechanisms. Dorzagliatin directly enhances the glucose-sensing capability of glucokinase, whereas liraglutide amplifies insulin secretion in response to glucose.

**Figure 6.**
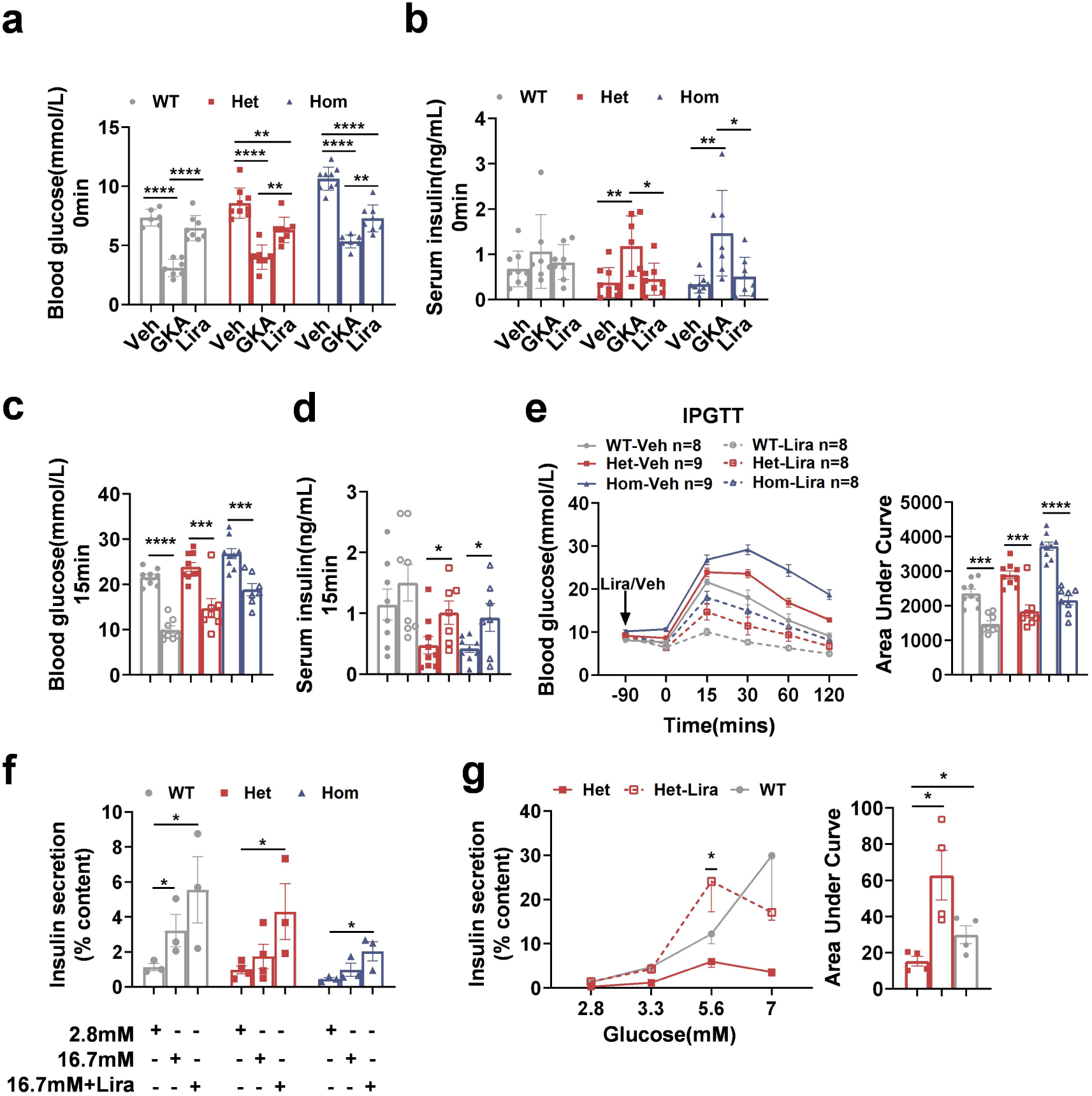
Liraglutide amplifies insulin secretion response to glucose and ameliorates glucose tolerance in GCK-Q26L mice. (**a, b**) For liraglutide, we conducted experiments similar to those shown in Figure 5 with dorzagliatin, assessing blood glucose (**a**) and serum insulin levels (**b**) in mice that had been treated with dorzagliatin and liraglutide prior to glucose challenge. (**c-e**) show the results of IPGTT with Liraglutide. Blood glucose (**c**) and serum insulin level (**d**) 15 minutes after glucose challenge. (**e**) IPGTT and the AUC of blood glucose. (**f**) Insulin secretion measured in isolated islets from 10-week-old male WT, Het and Hom mice when incubated in 2.8, 16.7 mmol/L glucose and 16.7 mmol/L glucose containing 100 nM Lira. (**g**) Dose-dependent effects of glucose on insulin secretion and the AUC of 10-week-old male WT and Het mice with or without 100 nM Lira, Het *vs* Het-Lira, * P < 0.05. Each data point represents an individual mouse. Values are shown as mean ± SEM. * P < 0.05 *** P < 0.001, **** P < 0.0001.

### A high PRS for insulin resistance is associated with atypical and more severe diabetic phenotypes in proband

The experimental evidence above demonstrates that GCK-Q26L is a GCK-MODY causing mutation. However, although the proband inherited the mutation from her father who presented typical clinical features of GCK-MODY, the proband presented more severe hyperglycemia that did not resemble a classic GCK-MODY (Fig. 1 and Table 1), suggesting that additional factors may contribute to phenotypes of the proband. Indeed, as the disease progressed, the proband became overweight (BMI 26.76 kg/m^2^) and developed dyslipidemia with a high Triglyceride (TG) to HDL-cholesterol (HDL-c) ratio, conditions often associated with insulin resistance [19]. Utilizing data from 424,068 individuals in the UK Biobank (UKB), we examined the association between the TG/HDL-c ratio, a surrogate measure of insulin resistance [9, 16–19], with insulin resistance polygenic scores. Results indicated the PRS was significantly associated with the observed TG/HDL-c ratio, that higher PRS levels corresponded with increases in the TG/HDL-c ratio, supporting the relevance of PRS in assessing insulin resistance genetic risk in the population (Fig. 7a). To explore the genetic predisposition of insulin resistance in this family, we conducted a PRS analysis and used the East Asian samples from the 1000 Genomes Project samples for evaluate the PRS risk and adjusted for potential population structure confounding through principal component analysis (PCA) (ESM Fig. 5a, b). The PRS analysis of the family indicated the proband inherited a high insulin resistance genetic risk from her mother, who has an IR PRS score ranked in the top 2% in the study population (Fig. 7b). According to previous studies, monogenic risk and polygenic scores may jointly influence the risk of disease [14]. These data suggest that the probands’ atypical and severe diabetic phenotypes were associated with high PRS for insulin resistance in addition to GCK mutation.

**Figure 7.**
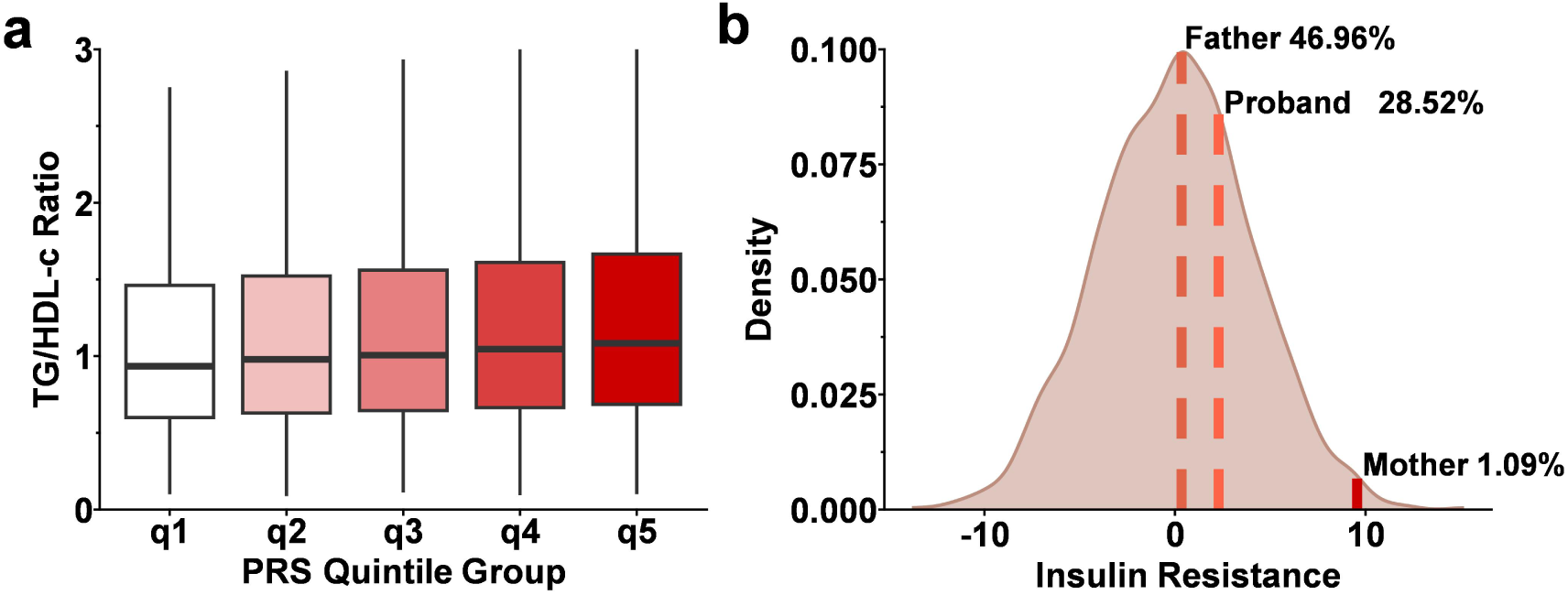
Quality control and PRS score for study samples. (**a**) The distribution of the ratio of triglycerides to HDL cholesterol as a function of quintiles of an insulin resistance (IR) polygenic risk score (PRS) was evaluated in 424,664 UK Biobank samples. The horizontal line in the box indicates the median value, and the top and bottom of the box indicate the upper and lower quartiles, respectively. (**b**) The results of insulin resistance polygenic risk score analysis on case family. The curve denotes the probability density function of the IR PRS scores from the 1000 Genomes Project East Asian samples. Vertical dashed lines indicate the PRS for each family member: the father’s position at the top 46.96^th^ percentile, the proband at the top 28.52^th^ percentile, and the mother at the top 1.09^th^ percentile.

## Discussion

The identification of the novel GCK-Q26L mutation, originally classified as VUS, in two members of a family with marked differences in PRS for insulin resistance and different severities of diabetes adds to the growing complexities of genetic contributions to the development and progression of diabetes. With multiple independent approaches, we demonstrated that GCK-Q26L did not affect insulin biosynthesis and islet insulin content, but impeded glucose sensing of pancreatic β cells, increasing the threshold of glucose concentration for insulin secretion, impairing GSIS, and therefore causing mild elevated FBG and glucose intolerance. These data established that GCK-Q26L is indeed a pathogenic mutation causing GCK-MODY.

GCK-MODY typically presents with mild hyperglycemia that is often managed without pharmacological intervention due to low risks of diabetes related chronic complications and the limited efficacy of traditional glucose-lowering medications [4, 6]. However, the presence of insulin resistance can complicate the clinical picture, leading to more severe hyperglycemia and necessitating therapeutic intervention. The results from our GCK-Q26L mouse model highlight the potential therapeutic benefits of using GKA and GLP-1RAs in managing diabetes phenotypes influenced by both GCK mutations and polygenic risk factors for insulin resistance. Dorzagliatin is a new a dual-acting GKA that enhances glucokinase activity by binding to its allosteric site, increasing glucose phosphorylation and insulin secretion, thereby improving glycemic control [23, 25]. In a clinical trial, dorzagliatin directly increases insulin secretion and glucose sensitivity in patients with GCK-MODY, suggesting that it may be able to preserve GCK activity in GCK-MODY [26]. Our data further demonstrated that dorzagliatin restored glucose threshold for insulin secretion and improved GSIS and glucose intolerance in GCK-Q26L mutant mice (Fig, 5d-e). It is worthy to note that dorzagliatin can increase insulin secretion in GCK-Q26L mice and decrease blood glucose even in the fasting state with relatively low FBG in both WT and mutant mice (Fig. 5a-c). However, there is no increased risk of hypoglycemia in a phase 3 clinical trial dorzagliatin [23]. It remains to be determined whether this was caused by extra high dose of dorzagliatin (about 15 times higher dose using in mice in this study than recommend dose for patients with type 2 diabetes) or due to different responses of different species.

In addition to dorzagliatin that directly targets to GCK, GLP-1RAs may also be considered for the management of patients with GCK-MODY due to its beneficial effect on glucose-dependent insulin secretion. Indeed, our data demonstrated that liraglutide enhanced glucose sensing of mutant islets at physiological glucose concentrations and improved GSIS (Fig. 6). Furthermore, a single administration of liraglutide significantly decreased FBG in mutant mice without affecting FBG in WT mice and markedly improved glucose tolerance (Fig, 6). These beneficial effects on glucose sensing and insulin secretion, along with its role in promoting weight loss and improving insulin resistance, make liraglutide (and possibly other GLP-1RAs) an attractive option for managing GCK-MODY, particularly in patients with GCK-MODY and insulin resistance.

To date, monogenic mutation involved in more than 70 genes have been identified to cause monogenic diabetes. Some of the biggest challenges for early recognition and diagnosis of monogenic diabetes are highly variable for disease penetrance and clinical presentations in patients carrying different mutations in one gene or even the same mutation in the same gene. Accumulating evidence indicates that genetic and/or environmental factors may exacerbate or lessen the conditions, and many VUS can have significant pathogenic effects when combined with other genetic predispositions [4, 27]. In the current study, we found that the proband who carried a GCK-MODY causing mutation inherited from her father, also possessed a high PRS for insulin resistance that may have been inherited in part from her mother, whose PRS for insulin resistance was in top 1.09^th^ percentile (Fig. 7). The elevated PRS may contribute to proband’s atypical presentation including more severe hyperglycemia and dyslipidemia, supporting an additive effect that monogenic and polygenic factors may interact. These findings align with recent research indicating that PRS can substantially influence the clinical presentation of monogenic diseases, including diabetes [28, 29]. This dual influence underscores the complexity of diabetes pathogenesis and challenges the traditional dichotomy between monogenic and polygenic diabetes, highlighting the importance of considering polygenic factors even in cases initially attributed to single-gene mutations [30, 31]. Incorporating comprehensive genetic assessments, including PRS, into routine diabetes care could revolutionize how we predict, diagnose, and manage diabetes. Recognizing the role of PRS might help identify individuals at high risk for severe forms of diabetes, facilitating earlier interventions and more personalized management strategies [32]. Additionally, understanding the interplay between different genetic factors could lead to the development of targeted therapies that address specific mechanistic pathways involved in the disease [33, 34]. Recent studies have demonstrated the potential of such personalized approaches in improving disease prognosis [35].

Despite these promising findings, several limitations need to be acknowledged. First, the study’s reliance on a mouse model may not fully capture the complexity of human GCK-MODY and its interaction with insulin resistance. While the GCK-Q26L mutant mouse model provides valuable insights, human clinical trials are essential to validate the efficacy and safety of GKA and GLP-1RAs in GCK-MODY individuals with or without insulin resistance. Second, the study did not explore the long-term effects of these treatments on beta-cell function and overall metabolic health. Longitudinal studies are necessary to understand the potential benefits and risks associated with chronic use of these therapies. Third, the sample size and genetic diversity of the study population were limited. Future research should include larger, more diverse cohorts to ensure the generalizability of the findings. Furthermore, exploring gene-environment interactions will be crucial in elucidating the full impact of monogenic and polygenic factors on the development and progression of monogenic diabetes.

## Author contributions

SJ, HS, and HZ designed and conducted experiments and analyzed data; SJ, HS, HZ YY, XL, SC, WF, JQ, JZ, and ZZ conducted experiments; All other authors revised the manuscript critically for important intellectual content. MW discussed project and edited manuscript; ML initiated and oversaw the project, designed the experiments, and wrote the manuscript.

## Supporting information

ESM

## Acknowledgments

The work was supported by the National Key R&D Program (2022YFE0131400 and 2019YFA0802502), and also by the National Natural Science Foundation of China (82220108014). We acknowledge the support of Tianjin Municipal Health Commission (TJWJ2021ZD001), Tianjin Medical University General Hospital Clinical Research Program (22ZYYLCZD02), Tianjin Municipal Science and Technology Commission (23JCQNJC00680 and 23JCQNJC00670), National Natural Science Foundation of China (82200892, 82301951, and 81800733), Tianjin Municipal Education Commission (2021KJ209), Tianjin Medical University Endocrine and Metabolic Disease the Youth Talent Incubator Program (2024XKNFM13), Tianjin Key Medical Discipline (Specialty) Construction Project (TJYXZDXK-030A), and Tianjin Medical University Clinical Special Disease Research Center - Neuroendocrine Tumor Clinical Special Disease Research Center.

## Data availability

Due to local privacy laws and privileged human information, all requests for raw genotyping and clinical data are subject to prior approval from the local IRB. Analysis of the UK Biobank data was performed using application 89885. The UK Biobank data are available to researchers with research inquiries following IRB and UK Biobank approval (https://www.ukbiobank.ac.uk/enableyour-research/apply-for-access).The polygenic scores described in this publication are available for download from the Polygenic Score Catalog (https://www.pgscatalog.org) under the publication ID PGP000223

## Code availability

The manuscript described a pragmatic approach of publicly available software. Additional code is available upon request from the authors.

## Notes

### Competing Interest Statement

The authors have declared no competing interest.

### Author Declarations

Tianjin Medical University General Hospital Ethics Committee gave ethical approval for this work

### Summary of Updates

Section on the first part results updated to clarify the frequency of the c.77A>T, pQ26L VUS in the general (normoglycemic) population and the pathogenicity predicted by the software.

