## Supplementary material for "Genotype-phenotype discrepancy among family members carrying a novel glucokinase mutation: insights into the interplay of GCK-MODY and insulin resistance": ESM

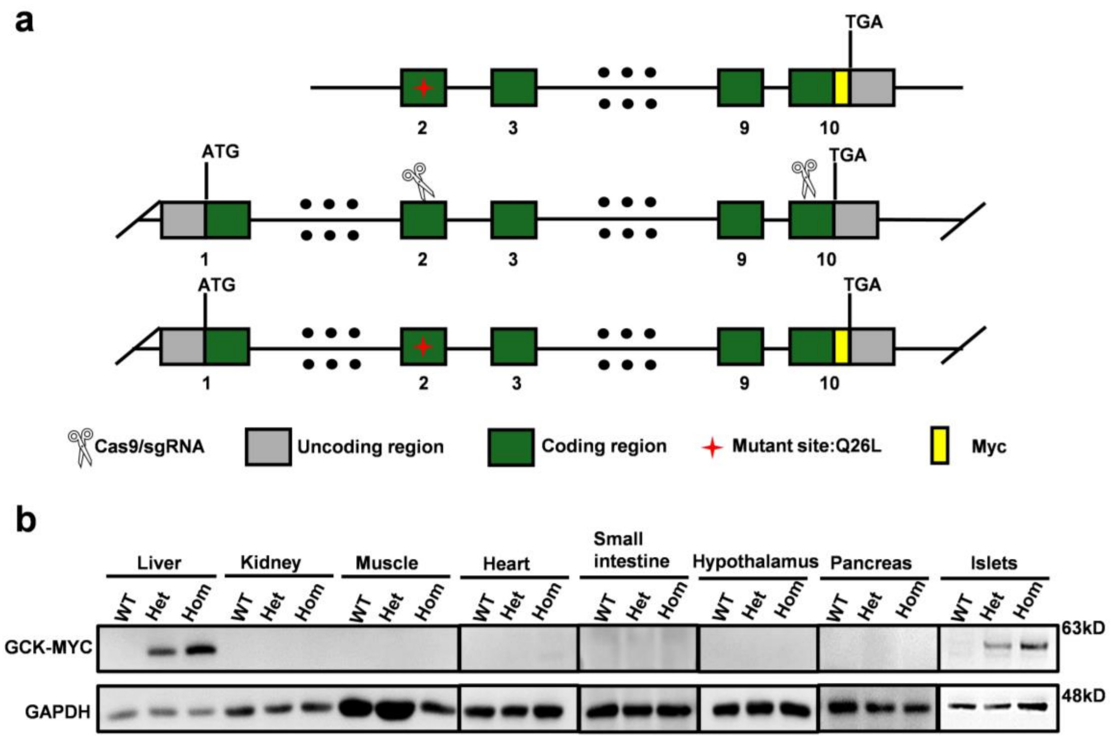


**ESM Figure 1. Construction and validation of a global GCK-Q26L knock-in mouse model.** (**a**) An A>T mutation at the 77$^{th}$ of the coding sequence of GCK gene was introduced using site-directed mutagenesis, resulting in a change of amino acid from glutamine (Q) to leucine (L) at GCK 26$^{th}$ residue. A Myc-tag was added just before the stop codon of GCK. The DNA fragment encoding Myc-tagged GCK-Q26L was then integrated into genome mediated by Crispr/Cas9. (**b**) Western blots examining the expression of Myc-tagged GCK in different tissues (liver, kidney, muscle, heart, hypothalamus, small intestine, hypothalamus, pancreas, and islets) using anti-Myc as indicated.

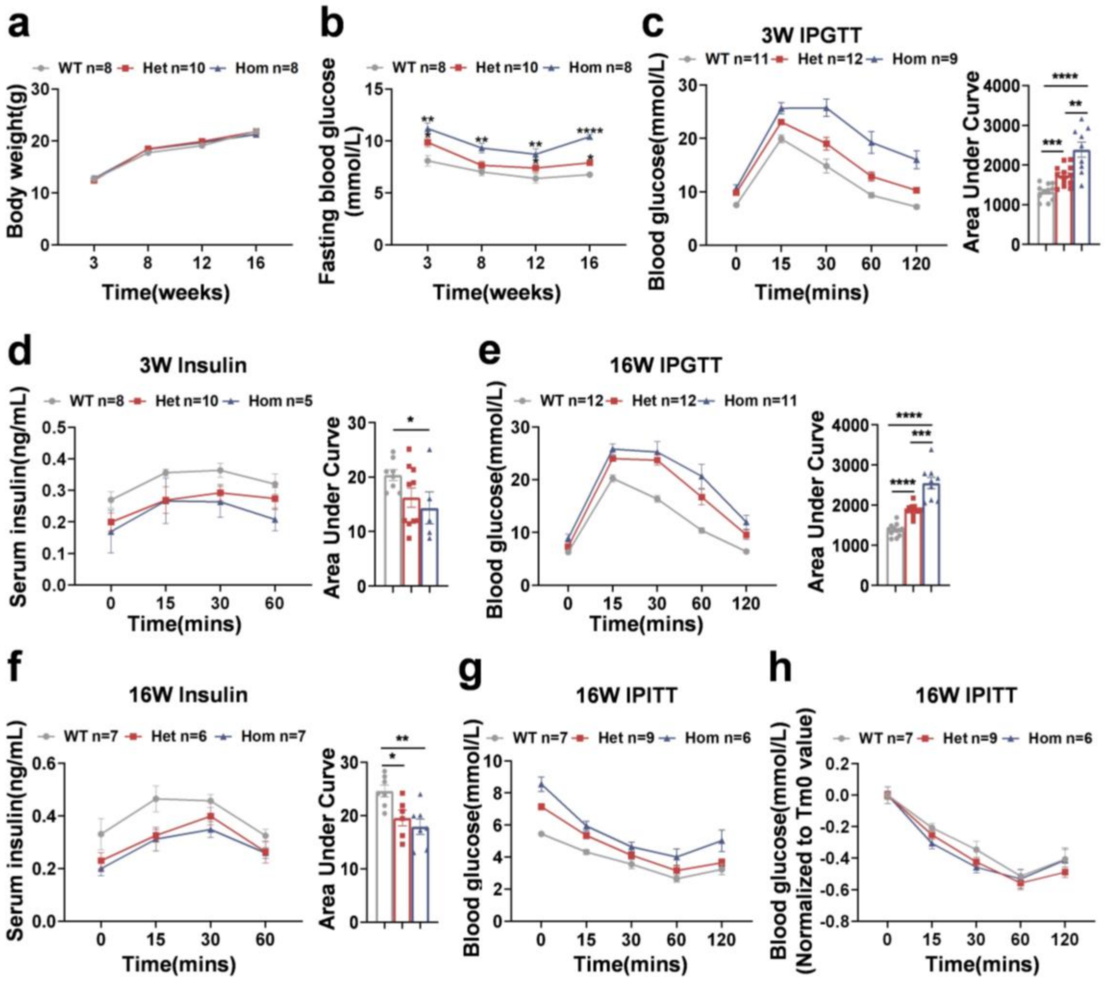


**ESM Figure 2. Diabetic phenotype in female mice.** (**a**,**b**) Body weight (**a**) and fasting blood glucose (**b**) of female mice were measured at 3, 8, 12, 16 weeks of age. WT *vs* Het, WT *vs* Hom, * P < 0.05, ** P < 0.01, **** P < 0.0001. (**c**) IPGTT in 3-week-old female mice (2 g/kg i.p.) and the area under curve (AUC) of blood glucose during IPGTT. (**d**) Serum insulin levels in 3-week-old female mice during IPGTT were measured by ELISA, with calculated AUCs. (**e**) IPGTT in 16-week-old female mice (2 g/kg i.p) and the AUC of blood glucose. (**f**) Serum insulin levels in 16-week-old female mice during IPGTT were measured by ELISA, with calculated AUCs. (**g**,**h**) IPITT were performed in 16-week-old female mice (0.75 U/kg i.p), (**h**) was (**g**) normalized to Tm0 value. Each data point represents an individual mouse. Values are shown as mean ± SEM. *P < 0.05, ** P < 0.01, *** P < 0.001, **** P < 0.0001.

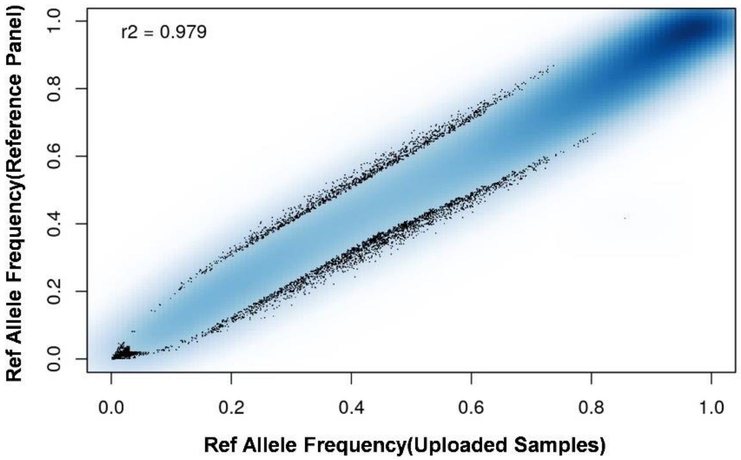


**ESM Figure 3. Correlation plots and the associated $R^2$ quality for ASA chip Case family mixed with 1000 Genomes samples with TOPMed reference panel.** The correlation coefficient $R^2$ is 0.979.

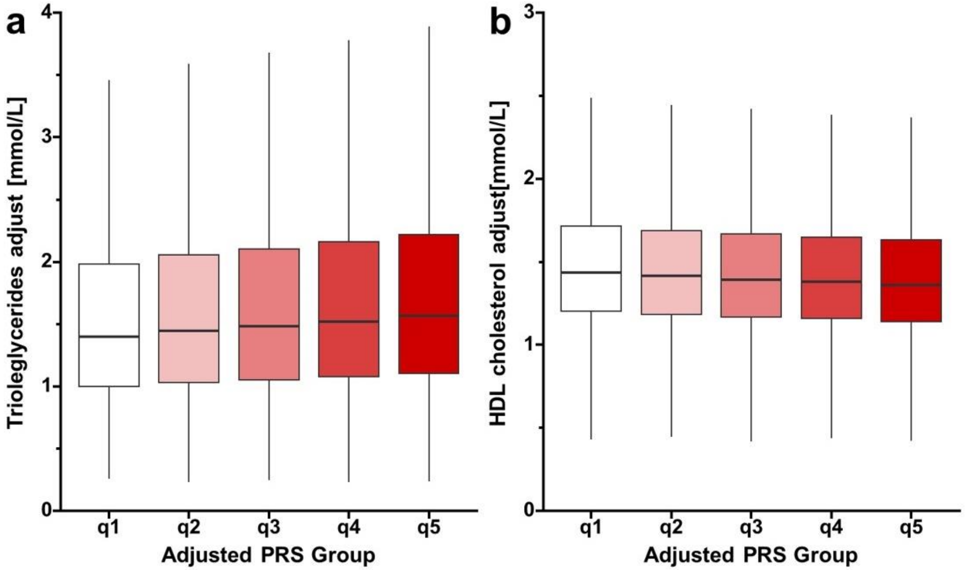


**ESM Figure 4. In the validation dataset, stratification according to polygenic score, box plots for Triglycerides adjust and HDL cholesterol adjust.** (a) Triglycerides adjust. (b) HDL cholesterol adjust.

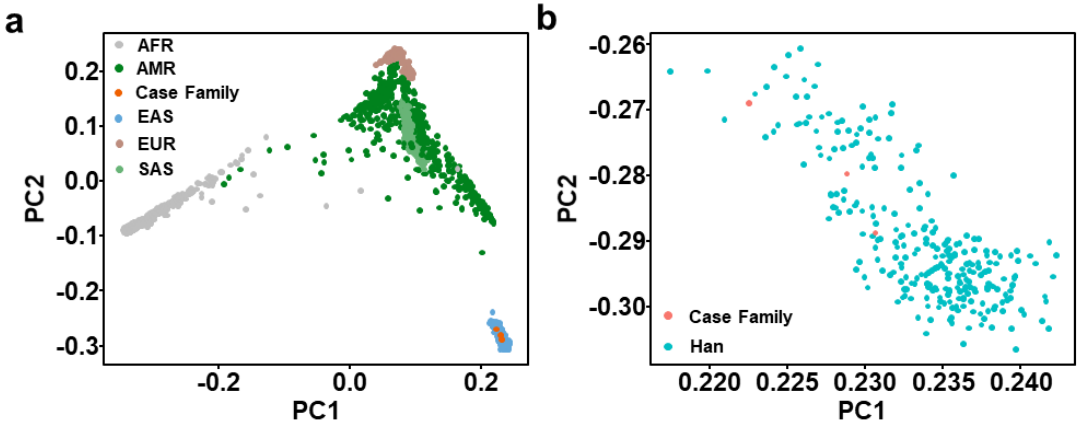


**ESM Figure5.** (a, b) PCA plot for case family mixed with 1000 Genomes Project samples. EUR, European; EAS, East Asian; AMR, admixed American; SAS, South Asian; African American; AFR, African.

**ESM table 1. Quality check for chip data**

| FID1 | IID1 | FID2 | IID2 | HETHET | IBS0 | KINSHIP |
|---|---|---|---|---|---|---|
| **CASE-CHIP** | CASE-CHIP | CASE-CHIP | F | 0.262585 | 0.00084542 | 0.265106 |
| **CASE-CHIP** | M | CASE-CHIP | F | 0.190004 | 0.067152 | 0.0524958 |
| **CASE-CHIP** | M | CASE-CHIP | CASE-CHIP | 0.25266 | 0.00110898 | 0.256758 |

Note: that KING kinship coefficients are scaled such that duplicate samples have kinship 0.5, not 1. First-degree relations (parent-child, full siblings) correspond to ~0.25, second-degree relations correspond to ~0.125, etc. It is conventional to use a cutoff of ~0.354 (the geometric mean of 0.5 and 0.25) to screen for monozygotic twins and duplicate samples, ~0.177 to add first-degree relations, etc. FID, family ID; IID, individual ID, HETHET, heterozygosity; IBS0, identity by State; KINSHIP, kinship coefficient.

**ESM table 2. Gender check for chip data**

| FID | IID | PEDSEX | SNPSEX | STATUS | F |
|---|---|---|---|---|---|
| **CASE-CHIP** | case | 2 | 2 | OK | -0.5504 |
| **CASE-CHIP** | case F | 1 | 1 | OK | 0.8417 |
| **CASE-CHIP** | case M | 2 | 2 | OK | -0.5746 |

Note: plink sex check on genetic chip data, including columns for family ID (FID), individual ID (IID), recorded gender (PEDSEX), gender inferred from SNP data (SNPSEX), status (STATUS), and a gender check statistic (F). table is used to confirm whether the gender recorded for samples matches the genetic data, thus helping to identify and correct potential errors in sample processing or data recording. The status "OK" indicates a match between recorded and genetic genders, while the F value provides a numerical basis for gender determination, with negative values typically indicating male and positive values indicating female.

**ESM table 3. Summary of the basic information used by UK Biobank to verify the data**

| | **Mean** | **Std. Dev** | **N total after excluding missing value** |
|---|---|---|---|
| **Age** | 56.55 | 8.09 | 424664 |
| **BMI (kg/m$^2$)** | 27.43 | 4.78 | 422958 |
| **HbA1c (mmol/L)** | 36.13(5.46%) | 6.78(0.62%) | 424664 |
| **TG (mmol/L)** | 1.81 | 1.09 | 467969 |
| **HDL-cholesterol (mmol/L)** | 1.45 | 0.38 | 424068 |
| **LDL-cholesterol (mmol/L)** | 3.77 | 0.86 | 397140 |
| **SBP (mmHg)** | 141.51 | 20.87 | 397128 |
| **DBP (mmHg)** | 83.39 | 11.44 | 404219 |

Note: Summarizes basic clinical and biochemical data utilized by the UK Biobank to verify participant information. It includes the mean, standard deviation, and total number of records after excluding missing values for various parameters.TG HDL LDL SBP DBP is adjusted value. BMI, body mass index; TG, triglycerides; HDL, high density lipoprotein; LDL, low density lipoprotein; SBP, systolic blood pressure; DBP, diastolic blood pressure

**Supplemental table 4. Summary of Adjustments for Lipid Levels Based on Lipid-Lowering Medications**

| Medication | Number of Subjects | Lipid Values Adjustment | | Reference(s) |
|---|---|---|---|---|
| | | HDL | Triglycerides | |
| **Statin** | 81964 | NA | -15% | Cholesterol Treatment Trialists' (CTT) Collaboration et al. Lancet. 2010 Nov 13;376(9753) |
| **Ezetimibe** | 2931 | NA | NA | 1- Sudhop T et al. Circulation 2002;106:1943-1948<br>2- Cannon CP et al. NEJM 2015;372:2387-97 |

| | | | | |
|---|---|---|---|---|
| | | | | 3- Zhao Z et al. Medicine (Baltimore). 2019 Feb;98(6) |
| **Bile Acid Sequestrant** | 196 | NA | NA | Lloyd-Jones et al. JACC 2017;70(14) |
| **Fibrate** | 1422 | 10% | -35% | Birjmohun RS et al. JACC 2005;45:185-97 |
| **Niacin** | 83 | 15% | -20% | Birjmohun RS et al. JACC 2005;45:185-97 |
| **Not specified** | 3929 | NA | -15% | Assumed statin |

Summarizes the adjustments made to HDL and triglyceride levels based on the type of lipid-lowering medication taken by subjects. The adjustments are necessary for accurate estimation of untreated lipid levels. The number of subjects for each medication type is listed, along with the percentage adjustments for HDL and triglycerides. References are provided for the source of each adjustment value
